# A new and efficient enrichment method for metagenomic sequencing of monkeypox virus

**DOI:** 10.1101/2022.07.29.22278145

**Authors:** Pablo Aja-Macaya, Soraya Rumbo-Feal, Margarita Poza, Angelina Cañizares, Juan A. Vallejo, Germán Bou

## Abstract

**Background:** The methodology described in previous literature for monkeypox virus (MPXV) sequencing shows low efficiency when using metagenomic approaches. The aim of the present study was to evaluate a new fine-tuned method for extraction and enrichment of genomic MPXV DNA using clinical samples and to compare it to a non-enrichment metagenomic approach.

**Results:** A new procedure that allows sample enrichment in MPXV DNA, avoiding wasting the sequencing quota in human DNA, was designed. This procedure consisted of host DNA depletion using a saponin/NaCl combination treatment and DNase. After typical quality control, samples using the enrichment method contained around 98 % of reads not classified as human DNA, while the non-enrichment protocol showed around 5-10 %. When reads not belonging to Orthopoxvirus were removed, enriched samples kept about 50 % of the original read counts, while non-enriched ones kept only 2-7 %.

**Conclusions:** Results showed a very significant improvement in sequencing efficiency, increasing the number of reads belonging to MPXV, the depth of coverage and the trustworthiness of the consensus sequences. This, in turn, would allow for more samples to be included in a single cartridge, reducing costs and time to diagnosis, which can be very important factors when dealing with a contagious disease.

## Background

Human monkeypox (hMPX) is a zoonosis disease originated in the jungles of Central and West Africa. This infectious disease was discovered in 1958 in two different monkey research colonies belonging to a Danish research institute [1]. It was described for first time in a child in the Democratic Republic of the Congo in 1970. Later in the 1970s, there were forty seven cases of human monkeypox in Central and West African Countries (Zaire, Nigeria, Liberia, Sierra Leone and Ivory Coast) [2]. In these areas the outbreaks of hMPX were reported in remote populations that depend on hunting and consume bushmeat [3]. Both rodents and monkeys can infect humans, however, it is not yet known which is the original reservoir of this disease [4].

The Global Commission for the Certification of Smallpox Eradication designated MPXV as the most important Orthopoxvirus infecting humans in the post-smallpox eradication era (from 1980). They recommended a surveillance program on MPXV and the study of its epidemiology and ecology [5].

Although the causes are unknown, since 1980 hMPX cases have gradually increased in Central Africa and more recently in West Africa [3]. In addition to this gradual increase in Africa, prior to 2022, hMPX cases outside of Africa were related to international travels or to animals imported from West and Central African countries [6]. However, from 2022, outbreaks with local transmission were established in multiple countries and continents [7]. Despite this increase, there is a lack of knowledge about hMPX emergence, epidemiology and ecology.

MPXV infects humans through contact with an infected animal, human or with contaminated material. MPXV enters the body through broken skin, the respiratory tract or the mucous membranes. Before the 2022 outbreaks, animals were the main transmission route for hMPX. This could occur by bite or scratch, bushmeat preparation, direct contact with body fluids, or lesions from an infected animal or contaminated material. However, 2022 outbreaks in different countries and continents showed that the main transmission route was human to human. This human to human transmission occurs by respiratory droplets, through contact with bodily fluids from infected people or with contaminated objects [7].

MPXV is an enveloped double-stranded DNA virus. This virus belongs to the Orthopoxvirus genus of the Poxviridae family and has a genome size of approximately 197 Kb. MPXV shares its genus with 11 species that affect different animals, such as the variola virus, which are historically important viruses [7]. Two clades of MPXV are currently distinguished by genomic sequencing: Central African and West African. Central African clade causes more severe disease and mortality [4]. MPXV and in general Poxviruses have excellent resistance to desiccation and wide pH tolerance compared with other enveloped viruses. These characteristics make the viral particles have a long stability in the environment. Materials from infected people or fomites could have infectious capacity during months or years. However, these viruses are sensitive to disinfectants, although less than others enveloped virus [7].

The aim of the present study was to evaluate and compare a metagenomic sequencing approach of MPXV that uses a regular DNA extraction (non-enrichment metagenomic approach) with a new MPXV DNA enrichment methodology proposed using clinical samples. The methodology described in the literature [8, 9, 10] showed a large waste of sequencing resources. For example, Cohen-Gihon et al. [8] obtained a total of 2 M sequences from MPXV sequencing, and only 48 K sequences belonged to MPXV (1.8 % of the total reads). Fuchs et al. [9] obtained 9 M reads from MPXV sequencing, and only 265 K reads belonged to MPXV (3 % of the total reads). In Israeli et al. [10] obtained 16.3 M reads from MPXV sequencing, and only 1 M reads belonged to MPXV (6.5 % of all reads). Isidro et al. [11] obtained around 80 million total reads *per* sample using a NextSeq 2000 (Illumina) device, but only 4 % of the reads belonged to MPXV. Overall, data indicates that over 90 % of the sequencing effort was wasted.

In this study, efforts were focused on improving the performance of the MPXV sequencing processes in order to avoid wasting most of the reads on the human host. This, in turn, allows for more samples to be included in the same sequencing run, which lowers costs and improves the coverage and the trustworthiness of the observed mutations.

## Results

A procedure that allows sample enrichment in MPXV DNA was designed, avoiding wasting the sequencing quota in human DNA. This procedure consisted of host DNA depletion using a saponin/NaCl combination treatment and DNase. Prior to this, a soft centrifugation that allowed the removal of big particles and part of the eukaryotic cells was used. After human DNA elimination, it is crucial to remove saponin, NaCl and DNase to generate a library for sequencing. In the present methodological proposal, samples were centrifuged at 35000 *g* and the MPXV particles washed three times using PBS. MPXV belongs to the Poxviridae family, characterized by being the most complex and largest viral family. This large size allowed their easy centrifugation at 35000 *g* [12].

MPXV samples used in this protocol are listed in Table S1. Two samples, MP01 and MP03 (anonymized identifiers), were processed using either the MPXV enrichment protocol proposed in this work (MP01CHUAC, MP03CHUAC) or the non-enrichment method (MP01bCHUAC, MP03bCHUAC). Sample groups were sequenced in two different runs, in order to generate a fair comparison.

Preliminary Kraken2 reports using the original reads (no filters or quality control), visualized in Figure 1, showed a clear difference between samples. In MP01CHUAC and MP03CHUAC there were almost no reads classified as host contamination, whereas in MP01bCHUAC and MP03bCHUAC most of the reads belonged to human DNA.

**Figure 1.**
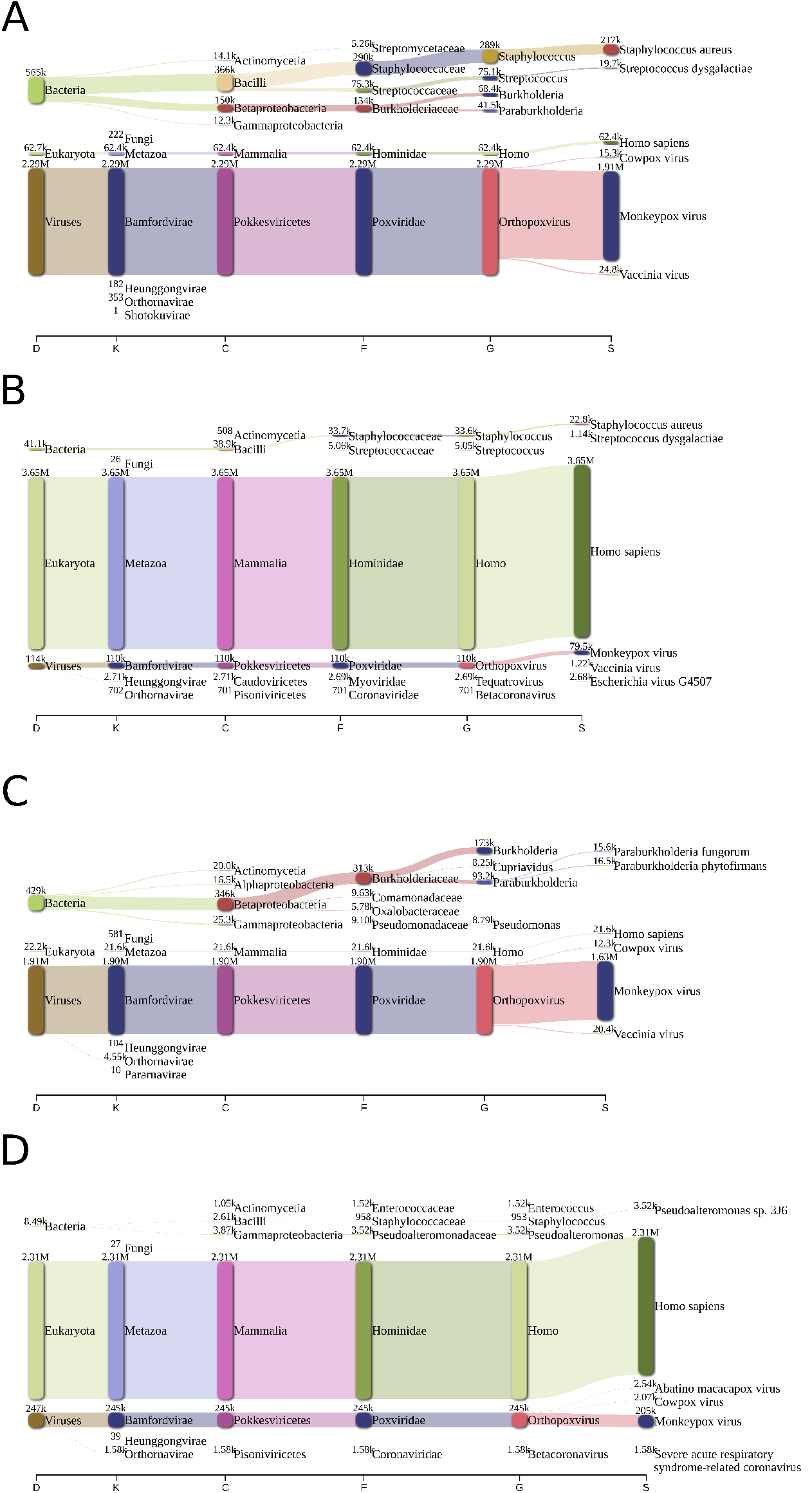
Raw read classification: These plots represent the classification Kraken2 has performed on the original reads (those without any filtering or quality control). The interpretation must be done with caution, as a lot of the hits are false positives, although the main ones, like MPXV, *Homo sapiens* or some bacteria, such as *Staphylococcus aureus*, are real. The read counts are presented in pairs and the maximum taxa *per* level is 6. A: MP01CHUAC, B: MP01bCHUAC, C: MP03CHUAC, D: MP03bCHUAC

In Figure 2, total read counts for each sample (including both forward and reverse reads) are presented for each quality control step. A significant change can be observed in the third step, BMTagger, were reads classified as human were removed. While read counts for MP01CHUAC and MP03CHUAC decreased from 4.75 M and 4.03 M reads to 4.65 M and 4.00 M, respectively (reduction of 1-2 %), the read counts for MP01bCHUAC and MP03bCHUAC went from 5.9 M and 4.02 M to 0.3 M and 0.4 M, respectively (reduction of 90-95 %). In the fourth step, Kraken2, where anything not classified as Orthopoxvirus is discarded, the differences were less drastic, with MP01CHUAC and MP03CHUAC having a reduction of 30 %, while MP01bCHUAC and MP03bCHUAC showed a reduction of 41 % and 8 %, respectively. When comparing the original read count with the final quality controlled reads, a reduction of about 50 % was observed for MP01CHUAC and MP03CHUAC, while for MP01bCHUAC and MP03bCHUAC it was around 93-98 %. The alignment statistics of the remaining reads against the reference genome “MPXV_USA_2022_MA001” (ON563414.3) are presented in Table 2, where MP01CHUAC and MP03CHUAC have a median depth of 1500-1800, while MP01bCHUAC and MP03bCHUAC have around 80-100.

**Table 1.** Read counts per quality control step. Read counts, shown for each quality control step, are calculated taking into account forward and reverse sequences separately. MP01CHUAC and MP03CHUAC followed the described enrichment protocol, while MP01bCHUAC and MP03bCHUAC followed a non-enrichment method. Original: raw reads; Illumina QC: Typical paired-end Illumina quality control; BMTagger: Human contamination removal; Kraken2: Removal of anything except MPXV.

| Sample | Original | Illumina QC | BMTagger | Kraken2 | Lost reads (%) |
| --- | --- | --- | --- | --- | --- |
| MP01CHUAC | 6,490,118 | 4,745,690 | 4,652,024 | 3,360,370 | 48.22 |
| MP01bCHUAC | 7,755,402 | 5,897,356 | 306,422 | 179,086 | 97.69 |
| MP03CHUAC | 5,702,680 | 4,030,170 | 4,002,176 | 2,695,288 | 52.74 |
| MP03bCHUAC | 5,204,934 | 4,020,926 | 406,616 | 374,718 | 92.80 |

**Table 2.**
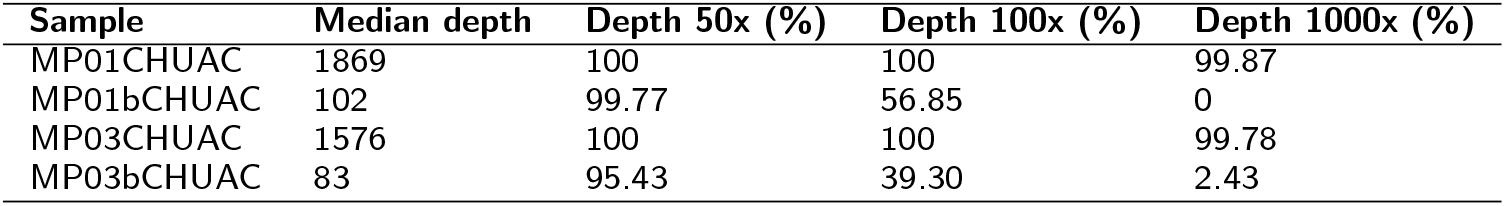
Alignment statistics per sample. Median depth and percentage of the genome with specific depths at various points is shown for each sample using the final quality controlled reads. MP01CHUAC and MP03CHUAC followed the described enrichment protocol, while MP01bCHUAC and MP03bCHUAC followed a non-enrichment method.

**Figure 2.**
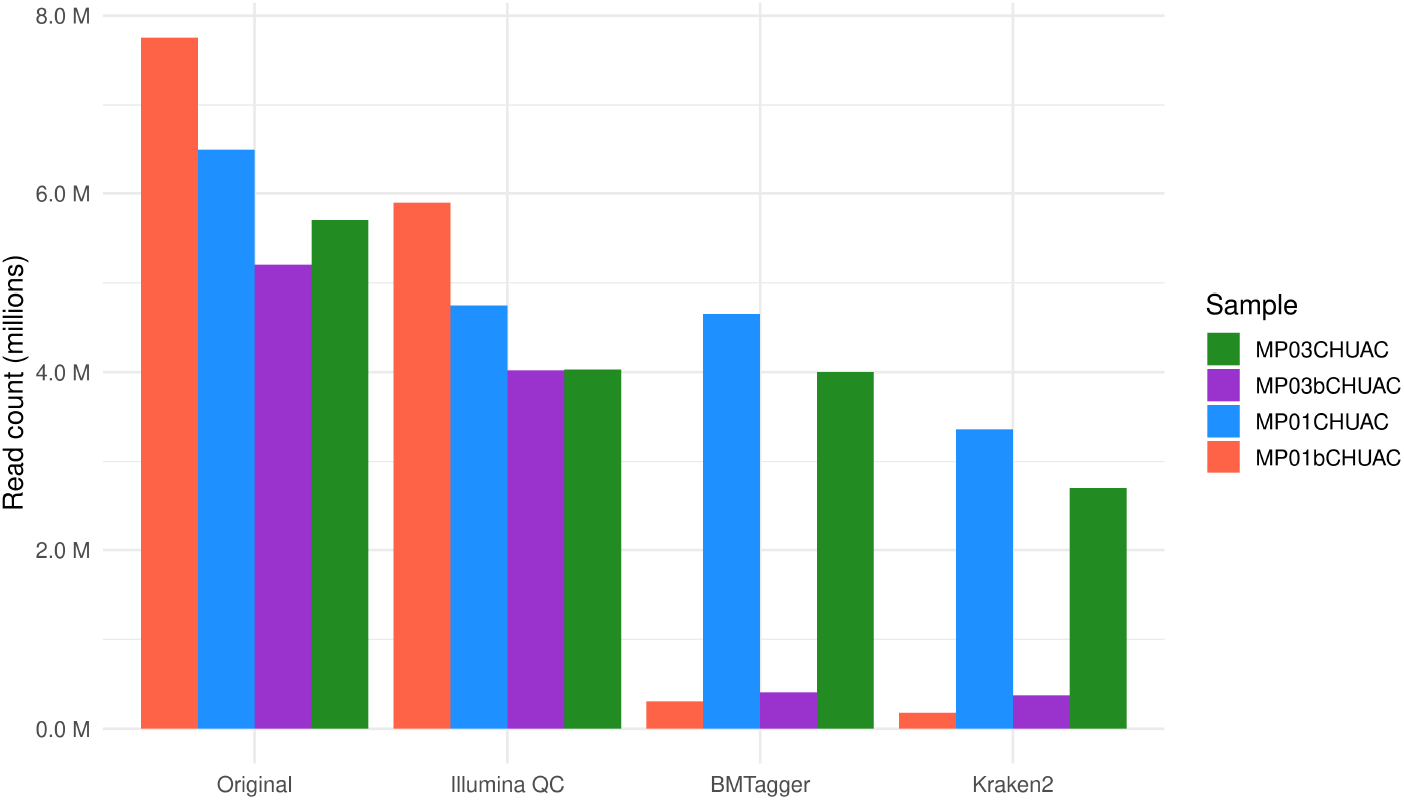
Read counts evolution across quality controls. The number of reads (counting both forward and reverse) is shown for each sample across various quality control steps. These steps are shown in order of execution, from left to right. Original: raw reads; Illumina QC: typical illumina quality control; BMTagger: removal of human reads; Kraken2: Removal of anything not belonging to the Orthopoxvirus genus.

All samples were able to produce a good quality consensus sequence belonging to lineage MPXV B.1 using an alignment-based consensus approach. Their SNPs (single nucleotide polymorphisms) against the reference genome “MPXV_USA_2022_MA001” (ON563414.3) are shown in Figure 3, with MP01CHUAC having a nucleotide substitution (G150330A) and MP03CHUAC having two aminoacid substitutions (OPG136:M389I, OPG163:H136Y). However, this approach may show problems in highly repetitive regions, such as the characteristic inverted terminal repeats (ITRs), and in some hot-spots containing very large and highly variable insertions. Due to this, a *de novo* assembly approach was also tested, which resulted in the same mutations and structure, but differing in the length of some insertions.

**Figure 3.**
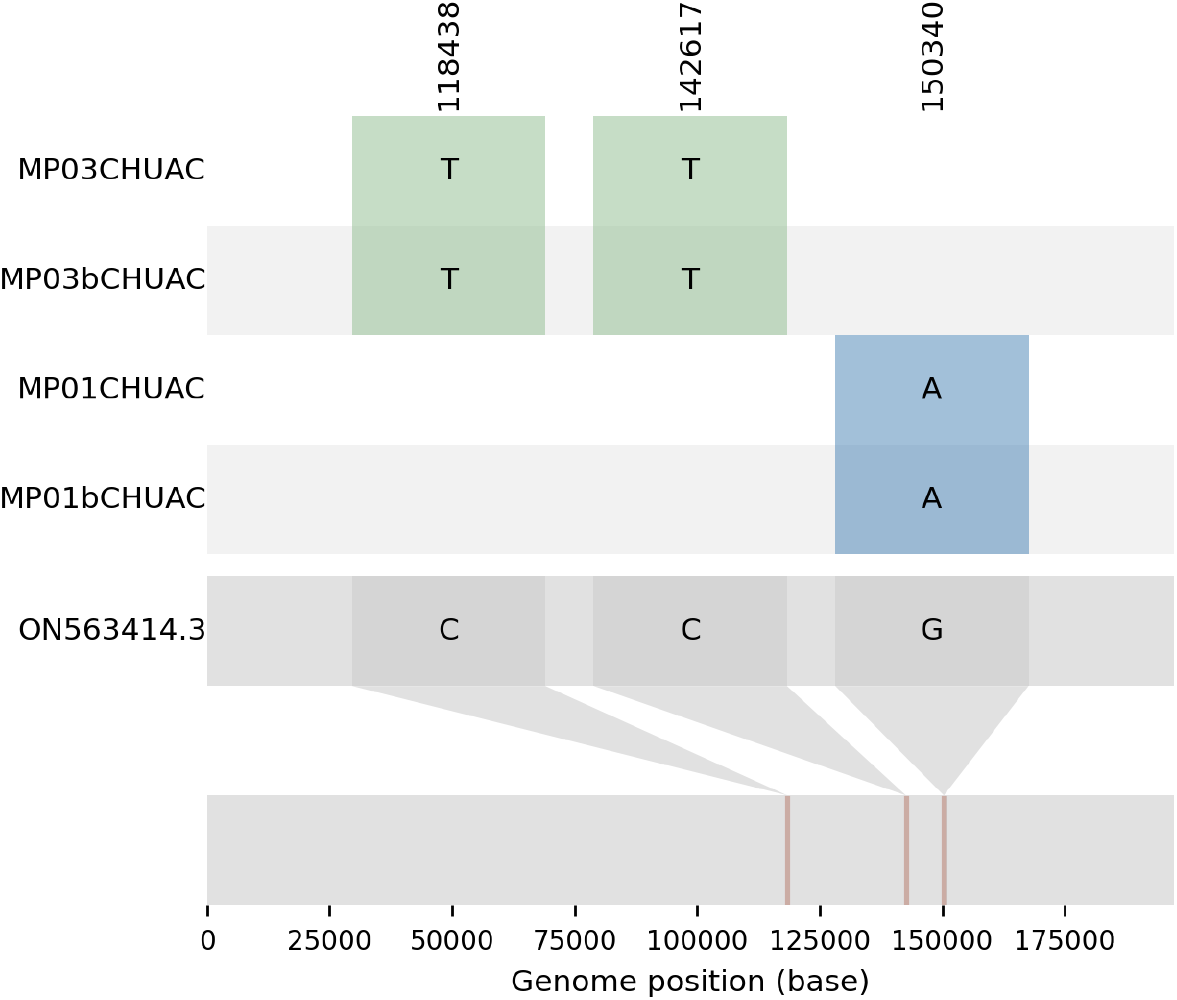
Substitutions detected in the MPXV used in this study: Single nucleotide polymorphisms (SNPs) for the samples used in this study are shown, comparing the mutations in each sample to the reference genome ON563414.3.

A phylogenomic analysis (Figure 4) was made to study the relatedness of the samples to all 275 complete MPXV genomes from taxid 10244 available in GenBank to date (2022-07-18, Table S2).

**Figure 4.**
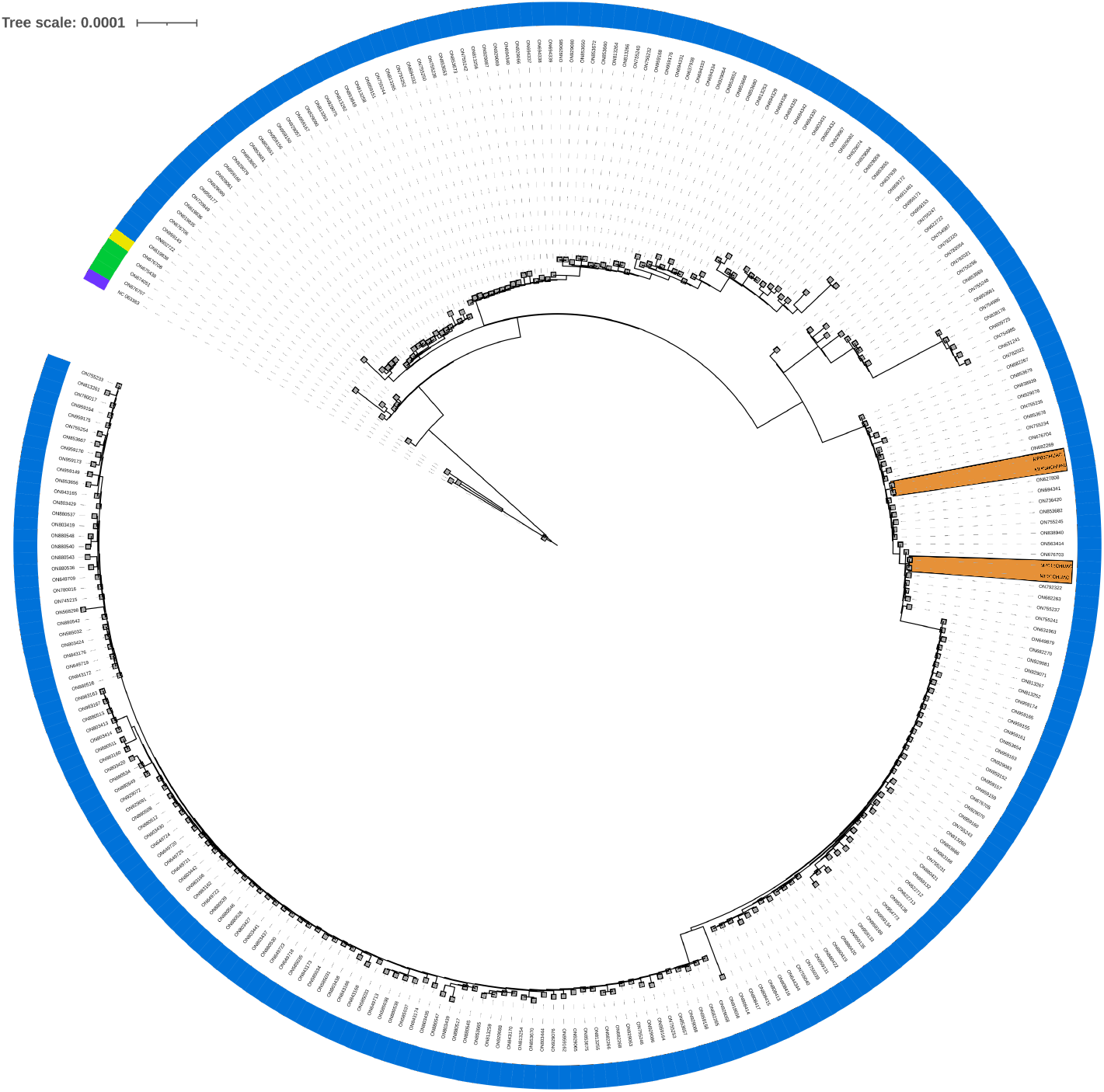
Phylogenomic tree. Phylogenomic analysis of the present study’s samples, comparing them to all complete MPXV genomes available in GenBank to date (2022-07-18, 275 genomes from taxid 10244). This does not pretend to infer the evolution of the virus, only to locate the most similar entries to the samples in this study. A color strip indicates each sample’s lineage (A: purple, A.1.1: yellow, A.2: green, B.1: blue), and orange areas highlight the study’s samples.

## Discussion

The objective of this study was to evaluate and compare a MPXV metagenomic sequencing method using a regular DNA extraction (non-enrichment approach) with a fine-tuned MPXV metagenomic sequencing method with MPXV DNA enrichment.

Results showed a significant differences when comparing depth and read count obtained using both methods. Specifically, the change in read count from the first quality control step (a typical procedure for any Illumina paired short reads) to the step where human reads are removed (Figure 2, step “BMTagger”), showed a reduction of 1-2 % for the enrichment protocol, whereas a reduction of 90-95 % was determined when using the non-enrichment protocol. Furthermore, when comparing the original reads with the final quality controlled reads, the MPXV DNA enrichment approach kept around 50 % of the reads, while the non-enrichment method kept 2-7 %. Additionally, the median depth when aligning the cleaned reads to the reference genome ON563414.3 was 1500-1800 for MP01CHUAC and MP03CHUAC and 80-100 for MP01bCHUAC and MP03bCHUAC.

When comparing alignment based consensus to *de novo* assemblies, the general structure was very similar (achieving a complete genome with both approaches), showing the same substitutions. However, hotspots where long and repetitive indels were detected caused problems in both methods, differing greatly in these areas. If the objective is to obtain a completely accurate and closed genome a hybrid approach should be utilized, using both short and long reads (e.g. Illumina and Oxford Nanopore Technologies). Nonetheless, characterizing these areas may not be as important for tracking the transmission of the disease. For example, Nextstrain’s pipeline for human monkeypox includes a step where masking of several regions of the genome is performed, including the first 1500 and last 7000 bp and repetitive regions of variable length.

All samples could produce good quality alignment based consensus, but the increased depth is one of the key elements to be able to trust the observed mutations. Furthermore, increasing the sequencing efficiency by removing human contamination before the sequencing procedure allows for more samples to be included in a single cartridge, which reduces costs and time to diagnosis, relevant when dealing with a contagious disease.

## Conclusions

Results showed a very significant improvement in sequencing efficiency, improving the number of reads belonging to MPXV, the depth of coverage and the trustworthiness of the consensus sequences. This, in turn, allows for more samples to be included in a single cartridge, reducing costs and time to diagnosis, which can be very important factors when dealing with a contagious disease.

## Methods

### Samples

Clinical samples were obtained from vesicular lesions or vesicular fluid swab and conserved in viral transport medium. Two samples that tested positive by qRT-PCR at the Microbiology Service of the A Coruña University Hospital (HUAC) were selected for this study. The remaining fractions of samples were stored at −20°C.

### DNA extraction

Viral DNA was extracted following two different protocols. In the first protocol (non-enrichment method), DNA extraction was performed using MagNA Pure Compact Nucleic Acid Isolation Kit I (Roche, Switzerland) following the manufacturer’s instructions and using 500 µL of viral transport medium as input. The second protocol (MPXV DNA enrichment) was designed to enrich samples in MPXV particles, modified from a saponine-based differential lysis method [13] followed by high g-force centrifugations. Briefly, 400 µL of samples were centrifuged at 10,000 *g* for 5 mins at 4°C. Supernatant was transferred to a tube for high g-force (Labcon, CA, USA) and centrifuged at 35,000 *g* for 30 min at 4°C. Pellet was resuspended in 250 µL of PBS supplemented with saponin 2.5 % and incubated at room temperature for 10 min. After the incubation, 350 µL of water were added and incubated for 30 s, and 12 µL of NaCl 5 M were also added. Samples were centrifuged at 35,000 *g* for 30 min at 4°C, pellets were resuspended in 100 µL of PBS and then 100 µL of NaCl 1 M, MgCl2 100 mM and 10 µL of HL-SAN DNase (ArticZymes, Norway Technologies) were added. Samples were incubated at 37°C for 15 min with shaking at 600 rpm. Following the incubation, samples were washed twice with 800 µL and 1 mL of PBS and centrifuged at 35,000 *g* for 30 min at 4°C after each wash. Final pellet was resuspended in 100 µL of nuclease-free water and nucleic acids were extracted using the QIAamp MinElute Virus Spin Kit (Qiagen, Germany) following the manufacturer’s instructions. DNA quantification was performed using the Qubit dsDNA HS Assay Kit (Thermo Fisher Scientific, MA, USA).

### Library generation and sequencing

DNA prep paired-end libraries (Illumina, CA, USA) were prepared using 1-5 ng of DNA extracted following the manufacturer protocol, except for the number of PCR cycles (15). Libraries concentration was quantified using the Qubit dsDNA HS Assay Kit (Thermo Fisher Scientific, MA, USA). DNA quality and fragment size of the libraries were evaluated using the High Sensitivity D1000 Kit for TapeStation 4150 (Agilent, CA, USA). Libraries were sequenced using a MiSeq platform (Illumina, CA, USA) using paired-end sequencing, with a read length of 150 nucleotides, using a V2 micro cartridge (Illumina, CA, USA) for every 2 samples.

### Bioinformatic analysis

Illumina reads were first processed using BBDuk (v. 38.96) [14] to remove PhiX contamination. Clumpify (v. 38.96) [14] was used to remove duplicates and to losslessly compress the files to minify space on disk. Finally, reads were trimmed with Trimmomatic (v. 0.39) [15] for adapter removal and quality control. Human contamination was removed using BMTagger [16]. Kraken2 (v. 2.1.2) [17] was used to classify reads using the full standard database (human, bacteria, plasmid, archaea, virus, fungi and UniVec_Core), extracting read count statistics and eliminating those that did not belong to the Orthopoxvirus genus. Visualization of these steps was facilitated by Pavian (v. 1.2.0) [18] and KrakenTools [17]. Other measures, such as read count at each quality control step were calculated with seqkit (v. 2.1.0) [19] Read quality was assessed before and after the entire cleaning process with FastQC (v. 0.11.9) [20] and MultiQC (v. 1.11) [21].

Reads that passed all the filters were aligned to the reference genome “MPXV_USA_2022_MA001” (ON563414.3) using BWA (v. 0.7.17-r1188) [22]. Duplicates were then marked with Picard (v. 2.27.4) [23] and alignment statistics (coverage, depth, aligned reads…) were calculated with BBMap’s pileup module (v. 38.96) [14]. A consensus sequence was then produced using iVar (v. 1.3.1) [24] (parameters: “-q 20 -t 0.5 -m 10”).

The alignment based consensus method was also compared to a *de novo* assembly approach, mainly due to the possible large indels that occur in MPXV. However, due to the large inverted terminal repeats (ITRs) in MPXV, *de novo* assemblies using a short read strategy can fail to represent both of these repetitive areas at the same time. In order to solve this, random subsets of N reads are made, which create different assemblies that when merged together can create a good scaffold, which is then polished using reads. This method can also utilize different assemblers and is inspired by Tricycler’s approach [25]. Possible drawbacks include: requiring high coverage and large repetitive insertions not being resolved with only short reads.

Various assemblies were created for each sample with Unicycler (v. 0.5.0) [26] (parameters: “–linear 1”) and SPAdes (v. 3.15.4) (parameters: “–trusted-contigs $ref -k 31,51,71”) [27] using subsets of reads chosen randomly by seqtk (v. 1.3-r106) [28]. The different assemblies were then aligned to the reference MPXV genome ON563414.3 with minimap2 (v. 2.24-r1122) [29], and a consensus was made using samtools (v. 1.15) [30]. Finally, the consensus was polished with Polypolish (v. 0.5.0) [31], the reads were aligned back to the polished genome and another consensus was made with iVar (v. 1.3.1).

Nextclade (v. 2.3.0) [32] was used to denote the lineages and to quickly visualize the quality of the sequences and their mutations. An SNP comparison was also created with snipit [33]. A more in depth phylogenomic analysis was made to see the relatedness of the study’s samples to all 275 complete MPXV genomes from taxid 10244 in GenBank to date (2022-07-18, Table S2). Sequences were aligned with Mafft (v. 7.453) [34] (parameters: “–auto”) to create a FASTA alignment, which was then transformed into PHYLIP format and used as input to RAxML (v. 8.2.12) [35] (parameters: “-m GTRCAT -T 60 -n tree -p 1 -N 1000 -p 12345 -x 12345 -f a”). The resulting tree was visualized with the Interactive Tree of Life (iTOL, v.6.5.8) [36].

## Supporting information

Samples used in this study

References used in phylogenomic tree

## Data Availability

All data produced in the present study are available upon reasonable request to the authors

## Acknowledgements

We thank the Microbiology personnel of the University Hospital of A Coruña for supplying the samples.

## Abbreviations

ITRs: Inverted terminal repeats
SNPs: Single nucleotide polymorphisms
hMPX: Human monkeypox
MPXV: Monkeypox virus

## Availability of data and materials

Consensus sequences for MP01CHUAC and MP03CHUAC are available under bioproject PRJNA863094. Cleaned reads containing only reads from monkeypox for all 4 samples are available under the same bioproject.

## Ethics approval and consent to participate

In order to satisfy any ethical or legal consideration, the study was carried out adhering to the standards of good clinical practice and current research regulations included in Law of Biomedical Research 14/2007, in accordance with the principles derived from the latest version of the Declaration of Helsinki and of the Convention on Human Rights and Biomedicine (the Oviedo Convention). Compliance with the protection of personal data of all those involved in the Organic Law 15/1999 and its implementing regulations, Royal Decree 1720/2007, is enforced. Sample names have been anonymized.

Additionally, the investigation ethics committee of A Coruña - Ferrol (CEI) states that this study does not require ethical oversight. The obtained information comes from pathogenic organisms, not from the patient’s clinical data. Neither the Spanish legislation for data protection and biomedical investigation, nor the European one, indicate that this study has to go through the revision of an ethics committee.

## Consent for publication

Not applicable.

## Authors’ contributions

Conceptualization, G.B., J.A.V, S.R, M.P. and P.A.; methodology, J.A.V, S.R, M.P. and P.A.; software, P.A.; validation, G.B., J.A.V, S.R, M.P., A.C. and P.A.; formal analysis, G.B., J.A.V, S.R, M.P. and P.A..; investigation, J.A.V, S.R and P.A.; resources, G.B., J.A.V, S.R, A.C., M.P. and P.A..; writing—original draft preparation, P.A., S.R. and J.A.V.; writing—review and editing, G.B., J.A.V, S.R, M.P. and P.A..F.-V.; visualization, G.B., J.A.V, S.R and P.A.; supervision, G.B and J.A.V.; project administration, G.B. and J.A.V.; funding acquisition, G.B and M.P. All authors have read and agreed to the published version of the manuscript.

## Additional Files

Supplementary Table 1 — Samples used in this study

Two samples, MP01 and MP03 (anonymized identifiers) were used in this study, each treated with two different protocols. MP01CHUAC and MP03CHUAC samples were applied a MPXV DNA enrichment method, while MP01bCHUAC and MP03bCHUAC samples were applied a non-enrichment protocol.

Supplementary Table 2 — References used in phylogenomic tree

All 275 complete MPXV genomes from taxid 10244 in GenBank to date (2022-07-18) used in the phylogenomic analysis.

## References

1. Magnus, P.v., Andersen, E.K., Petersen, K.B., Birch-Andersen, A.: A pox-like disease in cynomolgus monkeys. Acta Pathologica Microbiologica Scandinavica 46(2), 156–176 (1959)

2. Breman, J.G., Steniowski, M., Zanotto, E., Gromyko, A., Arita, I., et al.: Human monkeypox, 1970-79. Bulletin of the World Health Organization 58(2), 165 (1980)

3. Vandenbogaert, M., Kwasiborski, A., Gonofio, E., Descorps-Declère, S., Selekon, B., Nkili Meyong, A.A., Ouilibona, R.S., Gessain, A., Manuguerra, J.-C., Caro, V., et al.: Nanopore sequencing of a monkeypox virus strain isolated from a pustular lesion in the central african republic. Scientific reports 12(1), 1–13 (2022)

4. Petersen, E., Kantele, A., Koopmans, M., Asogun, D., Yinka-Ogunleye, A., Ihekweazu, C., Zumla, A.: Human monkeypox: epidemiologic and clinical characteristics, diagnosis, and prevention. Infectious Disease Clinics 33(4), 1027–1043 (2019)

5. World Health Organization (WHO): The Global Eradication of Smallpox: Final Report of the Global Commision for the Certification of Smallpox Eradication (1980). http://apps.who.int/iris/bitstream/handle/10665/39253/a41438.pdf Accessed Accessed 13 Jul 2022

6. Centers for Disease Control and Prevention (CDC): About Monkeypox (2022). https://www.cdc.gov/poxvirus/monkeypox/about.html Accessed Accessed 13 Jul 2022

7. European Centre for Disease Prevention and Control (ECDC): Factsheet for health professionals on monkeypox (2022). https://www.ecdc.europa.eu/en/all-topics-z/monkeypox/factsheet-health-professionals Accessed Accessed 13 Jul 2022

8. Cohen-Gihon, I., Israeli, O., Shifman, O., Erez, N., Melamed, S., Paran, N., Beth-Din, A., Zvi, A.: Identification and whole-genome sequencing of a Monkeypox virus strain isolated in Israel. Microbiology Resource Announcements 9(10), 01524–19 (2020)

9. Fuchs, J.: Travel-associated Monkeypox virus genomes from two German patients and of a derived virus isolate all closely related to a US sequence, 2022 (2022). https://virological.org/t/travel-associated-monkeypox-virus-genomes-from-two-german-patients-and-of-a-derived-virus-isolate-all-closely-related-to-a-us-sequence-2022/844 Accessed Accessed 15 Jul 2022

10. inbarg: First Israeli whole-genome sequence of Monkeypox virus associated with the May 2022 multi country outbreak (2022). https://virological.org/t/first-israeli-whole-genome-sequence-of-monkeypox-virus-associated-with-the-may-2022-multi-country-outbreak/843 Accessed Accessed 15 Jul 2022

11. Isidro, J., Borges, V., Pinto, M., Sobral, D., Santos, J.D., Nunes, A., Mixãao, V., Ferreira, R., Santos, D., Duarte, S., et al.: Phylogenomic characterization and signs of microevolution in the 2022 multi-country outbreak of monkeypox virus. Nature Medicine, 1–1 (2022)

12. Zwartouw, H., Westwood, J., Appleyard, G.: Purification of pox viruses by density gradient centrifugation. Microbiology 29(3), 523–529 (1962)

13. Charalampous, T., Kay, G.L., Richardson, H., Aydin, A., Baldan, R., Jeanes, C., Rae, D., Grundy, S., Turner, D.J., Wain, J., et al.: Nanopore metagenomics enables rapid clinical diagnosis of bacterial lower respiratory infection. Nature biotechnology 37(7), 783–792 (2019)

14. Bushnell, B.: BBmap. http://sourceforge.net/projects/bbmap Accessed Accessed 15 Jul 2022

15. Bolger, A.M., Lohse, M., Usadel, B.: Trimmomatic: a flexible trimmer for Illumina sequence data. Bioinformatics 30(15), 2114–2120 (2014)

16. WestGrid: BMTagger. https://www.westgrid.ca/support/software/bmtagger Accessed Accessed 15 Jul 2022

17. Wood, D.E., Lu, J., Langmead, B.: Improved metagenomic analysis with Kraken 2. Genome biology 20(1), 1–13 (2019)

18. Breitwieser, F.P., Salzberg, S.L.: Pavian: interactive analysis of metagenomics data for microbiome studies and pathogen identification. Bioinformatics 36(4), 1303–1304 (2020)

19. Shen, W., Le, S., Li, Y., Hu, F.: SeqKit: a cross-platform and ultrafast toolkit for FASTA/Q file manipulation. PloS one 11(10), 0163962 (2016)

20. Andrews, S., et al.: FastQC: a quality control tool for high throughput sequence data. Babraham Bioinformatics, Babraham Institute, Cambridge, United Kingdom (2010)

21. Ewels, P., Magnusson, M., Lundin, S., Käller, M.: MultiQC: summarize analysis results for multiple tools and samples in a single report. Bioinformatics 32(19), 3047–3048 (2016)

22. Li, H.: Aligning sequence reads, clone sequences and assembly contigs with BWA-MEM. arXiv preprint arXiv:1303.3997 (2013)

23. Broad Institute: Picard. https://github.com/broadinstitute/picard Accessed Accessed 15 Jul 2022

24. Grubaugh, N.D., Gangavarapu, K., Quick, J., Matteson, N.L., De Jesus, J.G., Main, B.J., Tan, A.L., Paul, L.M., Brackney, D.E., Grewal, S., et al.: An amplicon-based sequencing framework for accurately measuring intrahost virus diversity using PrimalSeq and iVar. Genome biology 20(1), 1–19 (2019)

25. Wick, R.R., Judd, L.M., Cerdeira, L.T., Hawkey, J., Méric, G., Vezina, B., Wyres, K.L., Holt, K.E.: Trycycler: consensus long-read assemblies for bacterial genomes. Genome biology 22(1), 1–17 (2021)

26. Wick, R.R., Judd, L.M., Gorrie, C.L., Holt, K.E.: Unicycler: resolving bacterial genome assemblies from short and long sequencing reads. PLoS computational biology 13(6), 1005595 (2017)

27. Bankevich, A., Nurk, S., Antipov, D., Gurevich, A.A., Dvorkin, M., Kulikov, A.S., Lesin, V.M., Nikolenko, S.I., Pham, S., Prjibelski, A.D., et al.: SPAdes: a new genome assembly algorithm and its applications to single-cell sequencing. Journal of computational biology 19(5), 455–477 (2012)

28. Li, H.: seqtk. https://github.com/lh3/seqtk Accessed Accessed 15 Jul 2022

29. Li, H.: Minimap2: pairwise alignment for nucleotide sequences. Bioinformatics 34(18), 3094–3100 (2018)

30. Li, H., Handsaker, B., Wysoker, A., Fennell, T., Ruan, J., Homer, N., Marth, G., Abecasis, G., Durbin, R.: The sequence alignment/map format and samtools. Bioinformatics 25(16), 2078–2079 (2009)

31. Wick, R.R., Holt, K.E.: Polypolish: Short-read polishing of long-read bacterial genome assemblies. PLoS computational biology 18(1), 1009802 (2022)

32. Aksamentov, I., Roemer, C., Hodcroft, E.B., Neher, R.A.: Nextclade: clade assignment, mutation calling and quality control for viral genomes. Journal of Open Source Software 6(67), 3773 (2021)

33. aineniamh: snipit. https://github.com/aineniamh/snipit Accessed Accessed 15 Jul 2022

34. Katoh, K., Toh, H.: Recent developments in the MAFFT multiple sequence alignment program. Briefings in bioinformatics 9(4), 286–298 (2008)

35. Stamatakis, A.: RAxML version 8: a tool for phylogenetic analysis and post-analysis of large phylogenies. Bioinformatics 30(9), 1312–1313 (2014)

36. Letunic, I., Bork, P.: Interactive Tree Of Life (iTOL) v5: an online tool for phylogenetic tree display and annotation. Nucleic acids research 49(W1), 293–296 (2021)

